# Anemia and Immune-Hematologic profiles in Virally Suppressed People with HIV: Findings from a Cross-Sectional Study

**DOI:** 10.1101/2025.08.01.25332652

**Authors:** Kingsley Kamvuma, Benson M. Hamooya, Sody M Munsaka, Sepiso K. Masenga

## Abstract

**Background:** Anemia remains a prevalent complication among people living with HIV (PLWH), even among virally suppressed PLWH. Sustained Immune activation, erythropoietin deficiency and disturbances in iron metabolism are thought to contribute to persistent anemia, yet their roles remain poorly defined in this population. This study investigated immune-hematologic profiles associated with anemia in virally suppressed PLWH.

**Methods:** A cross-sectional study was conducted among 155 virally suppressed PLWH attending the Livingstone University Teaching Hospital. Participants were classified as anaemic or non-anaemic based on WHO haemoglobin criteria. Demographic, clinical, and laboratory data including cytokines, inflammatory markers, and iron metabolism indices were collected. Descriptive statistics, bivariate analyses, and logistic regression models were used to evaluate associations with anemia.

**Results:** Anemia was present in 28.4% (95% CI: 21.4%–36.4%) of participants and was significantly more common in females than males (40.9% vs. 15.6%, p = 0.002). In the adjusted logistic regression models, increasing age was significantly associated with higher odds of anemia (AOR = 1.13; 95% CI: 1.021–1.252; p = 0.018). Among the cytokines analyzed, interferon-gamma (IFN-γ) was the only marker significantly elevated in participants with anemia (AOR = 1.003; 95% CI: 1.001–1.005; p = 0.012), while interleukin-17a (IL-17a) demonstrated a borderline inverse association (AOR = 0.99; 95% CI: 0.99–1.00; p = 0.051). Among hematologic markers, a soluble transferrin receptor-to-ferritin (sTfR-Ferritin) index >2 was significantly associated with anemia (AOR = 6.54; 95% CI: 1.001–42.76; p = 0.048), alongside female sex (AOR = 10.01; 95% CI: 1.08–92.64; p = 0.042).

**Conclusion:** Anemia remains a prevalent comorbidity among virally suppressed PLWH, particularly in women. Independent associations with anemia include advancing age, elevated interferon-gamma levels, and an increased soluble transferrin receptor-to-ferritin index, reflecting immune activation and depleted iron stores. These findings underscore the need for integrated monitoring strategies beyond virologic control, incorporating immune and iron metabolism biomarkers to improve the early detection and management of anemia in this population.

## Introduction

Anemia remains a significant yet underappreciated comorbidity among people living with HIV (PLWH), despite viral suppression through effective antiretroviral therapy (ART) (1). The global prevalence of anemia in PLWH is alarmingly high, with estimates suggesting that it affects up to 90% of individuals, often contributing to poor health outcomes, diminished quality of life, and increased mortality rates (2). While the advent of ART has dramatically improved survival rates and viral control, the persistent burden of anemia (3) highlights the need for a deeper understanding of its underlying pathophysiology.

Anemia in HIV has been attributed to several factors, including opportunistic infections, bone marrow suppression due to ART, and deficiencies in micronutrients(4). Chronic inflammation, often marked by elevated cytokines can impair erythropoiesis by suppressing erythropoietin synthesis and inhibiting bone marrow function (4). Inflammation promotes increased hepcidin levels, which reduce iron availability by blocking its release from macrophages and intestinal absorption, leading to functional iron deficiency (5). Additionally, immune activation may alter hematopoietic stem cell dynamics and contribute to anemia through oxidative stress and shortened red blood cell lifespan (6). Together, these processes may contribute to the development and persistence of anemia.

Iron metabolism plays a central role in the development of anemia, particularly in the context of HIV (7). The soluble transferrin receptor (sTfR) to ferritin index has emerged as a valuable diagnostic tool to distinguish between iron deficiency anemia and anemia of inflammation (8). Elevated sTfR reflects increased cellular iron demand, while low ferritin typically indicates depleted iron stores; however, ferritin is also an acute phase reactant and can be elevated in inflammatory states, potentially masking true iron deficiency (9). Thus, the sTfR/ferritin ratio provides a more accurate assessment of functional iron status, especially in the presence of chronic inflammation (10).

Despite the success of ART in achieving viral suppression among PLWH, anemia remains a prevalent and clinically significant complication (11). Factors beyond viral replication including chronic immune activation, inflammation, and disruptions in iron metabolism may play critical roles. However, the biological mechanisms underlying anemia in virally suppressed individuals remain poorly understood, particularly in sub-Saharan Africa where the burden of anemia in HIV is disproportionately high and mechanistic studies are limited (2). Understanding these mechanisms is critical, not only for improving diagnostic accuracy but also for developing targeted interventions aimed at alleviating anemia and improving overall health outcomes for PLWH.

## Methods

### Study Design and Setting

This was a cross-sectional analytical study conducted at the University Teaching Hospital, a tertiary referral center for HIV care and treatment in Zambia. This study investigated immune-hematologic profiles associated with anemia in PLWH. Following informed consent virally suppresed PLWH attending the ART clinic at the LUTH were recruited. Viral suppression was defined as a viral load of less than 1,000 copies/ml, according to the Zambia Consolidated Guidelines for HIV Treatment and Prevention (12).

### Eligibility Criteria

Participants were aged 18 years or older, HIV-positive, and had either anemia (as defined) or no anemia. Participants were required to have complete clinical and laboratory data available from the smart care database. Participants were excluded if they had any haematological malignancies, evidence of a recent infections. Additionally, pregnant women and women with abornomal uterine bleeding were not included in the study.

### Operational Definitions

Anemia was defined as a haemoglobin level <13 g/dL for men and <12 g/dL for women, according to World Health Organization criteria (13). Biomarkers of interest included hepcidin, erythropoietin, haemoglobin, ferritin, and soluble transferrin receptor and several inflammatory makers which were assessed to evaluate their correlation with anemia status. The soluble transferrin receptor to ferritin index is a diagnostic tool that helps distinguish iron deficiency anemia from anemia of chronic disease by combining sTfR (which reflects iron demand) and ferritin (which indicates iron stores) (14). It is especially useful in inflammatory conditions where ferritin may be falsely elevated. sTfR to ferritin index is a composite marker that helps differentiate iron deficiency anemia from anemia of inflammation, with higher values indicating depleted iron stores or pure iron deficiency (15).

### Sample Size

For the immune-hematologic profiles study among virally suppressed PLHIV, the sample size was determined using ferritin values as a marker. The study aimed for a 95% confidence interval and 90% power. The mean ferritin levels were 228.7 μg/L for anemic individuals (Group 1) and 270.1 μg/L for non-anemic individuals (Group 2), with standard deviations of 56.48 and 39.21, respectively based on previous study done in PLWH (16). Based on this, the total sample size was set at 155, with 39 participants in Group 1 (anemic) and 116 in Group 2 (non-anemic). The openepi software was utilized to calculate the sample size (17).

### Data Sources

We utilized a structured interview questionnaires were utilized to collect participant information, while patient files supplemented the data with clinical histories and treatment records.

### Data Collection and Study Variables

Data collected was on demographic including age, sex. Haematological profiles such as haemoglobin, ferritin, hepcidin, erythropoietin, and soluble transferrin receptor were also assessed. Additionally, immune activation and inflammatory markers, including CD4+ count, white blood cell count (WBC), absolute lymphocyte count, monocytes, IL-6, IFN-gamma, IL-17, D-dimer, high-sensitivity C-reactive protein (hs-CRP), IL-5, IL-6, IFN, and IL-1, were measured. Blood samples were processed for biomarker analysis using ELISA kits and other specialized laboratory equipments including the Pentra C200 and XT1800i hematology analyser. The neutrophil-to-lymphocyte ratio (NLR) was calculated as absolute neutrophil count divided by absolute lymphocyte count. The systemic immune-inflammation index (SII) was computed as (neutrophil count × platelet count) / lymphocyte count (18). These indices served as integrated profiles of immune activation and systemic inflammation. For ART regimens Non-nucleoside reverse transcriptase inhibitor regimens included efavirenz or nevirapine combined with either abacavir or tenofovir-based backbones. Protease inhibitor regimens consisted of lopinavir or atazanavir boosted with ritonavir and combined with abacavir, zidovudine, or tenofovir-based combinations. Integrase inhibitor regimens included dolutegravir with tenofovir and lamivudine.

### Statistical Analysis

Data was analysed using SPSS version 22. Descriptive statistics were used to summarize the characteristics of the study population. Comparisons between anemic and non-anemic groups for categorical variables, such as age, sex and Stfr1-Ferritin index, were performed using the Chi-square test.. Continuous variables were compared using Student’s t-test or Mann-Whitney U test, depending on the normality of the data, as determined by the Shapiro-Wilk test. To assess the associations between clinical and inflammatory factors and anemia, unadjusted and adjusted logistic regression analyses were performed. The adjusted model controlled for age and sex. Statistical significance was defined as a p-value < 0.05.

### Ethical Approval

Ethical approval for this study was obtained from the University of Zambia Biomedical Research Ethics Committee (UNZABREC; Reference No. 4062-2023). All participants were aged 18 years and above who provided written informed consent prior to participation were recruited. Participant confidentiality was ensured by assigning unique study identifiers and removing any personally identifiable information from the dataset. All electronic data were stored on password-protected systems accessible only to the research team, while physical records were kept in locked cabinets. The study adhered to the Declaration of Helsinki and Good Clinical Practice (GCP) guidelines at all stages. Data was collected between 5^th^ February and 24^th^ June 2024 at Livingstone University Teaching Hospital (LUTH), Zambia.

## Results

### Descriptive characteristics

The study involved 155 participants, the majority of whom were women (110 participants, 71.0%). Nearly all participants (95.4%) were on INSTI-based antiretroviral therapy and all had achieved viral suppression. Among them, 37 (23.9%) were married, 11 (7.1%) were smokers, and 43 (27.7%) were employed (Table 1).

**Table 1.** Demographic and clinical characteristics associated with anemia in people living with HIV.

| Variables | Total | Anemia |  | p |
| --- | --- | --- | --- | --- |
|  |  | Yes (28.4%) | No (71.6%) |  |
| **Sex** |  |  |  | **0.002** |
| Male | 45 | 7 (15.6) | 38 (84.4) |  |
| Female | 110 | 45 (40.9) | 65 (59.1) |  |
| **Married** |  |  |  | **0.746** |
| No | 89 | 22 (24.7) | 67 (75.3) |  |
| Yes | 37 | 13 (35.1) | 24 (64.9) |  |
| **STFR-Ferritin Index** |  |  |  | **0.023** |
| >2 | 17 | 10 (58.8) | 17 (41.2) |  |
| 1-2 | 42 | 18 (42.9) | 24 (56.1) |  |
| <1 | 47 | 8 (17.0) | 39 (83.0) |  |
| **Employment** |  |  |  | **0.234** |
| No | 111 | 36 (32.4) | 75 (66.6) |  |
| Yes | 43 | 15 (34.9) | 28 (65.1) |  |
| **ART regimens** |  |  |  | **0.367** |
| NNRTI | 4 | 2 (50) | 2 (50) |  |
| INSTI | 148 | 52 (35.1) | 96 (64.9) |  |
| PIs | 3 | 0 (0) | 3 (100) |  |
| **Smoking** |  |  |  | **0.600** |
| No | 137 | 46 (33.6) | 91 (66.4) |  |
| Yes | 11 | 3 (27.3) | 8 (76.7) |  |

Table 1 also highlights significant associations between anemia status and certain participant characteristics. Anemia was significantly more common among women than men (p=0.002), affecting 40.9% of women (45 out of 110) compared to 15.6% of men (7 out of 45). Additionally, the soluble transferrin receptor-ferritin (STFR-Ferritin) index showed a significant association with anemia (p=0.023); participants with an index <2 had a markedly higher prevalence of anemia (58.8%) compared to those with an index <1 (17.0%).

### Clinical Characteristics by Anaemic status PLWH

Figure 1 illustrates baseline clinical characteristics among virally suppressed PLWH stratified by anemia status. Statistically significant differences were found in alanine aminotransferase (ALT) and uric acid levels. Individuals with anemia exhibited significantly lower ALT levels (p < 0.05) and higher uric acid concentrations (p < 0.05) compared to those without anemia figure 1. Conversely, there were no significant differences in other clinical parameters, including age, body mass index (BMI), estimated glomerular filtration rate (eGFR), or the duration on antiretroviral therapy (ART), as indicated by ‘ns’ (not significant).

**Figure 1.**
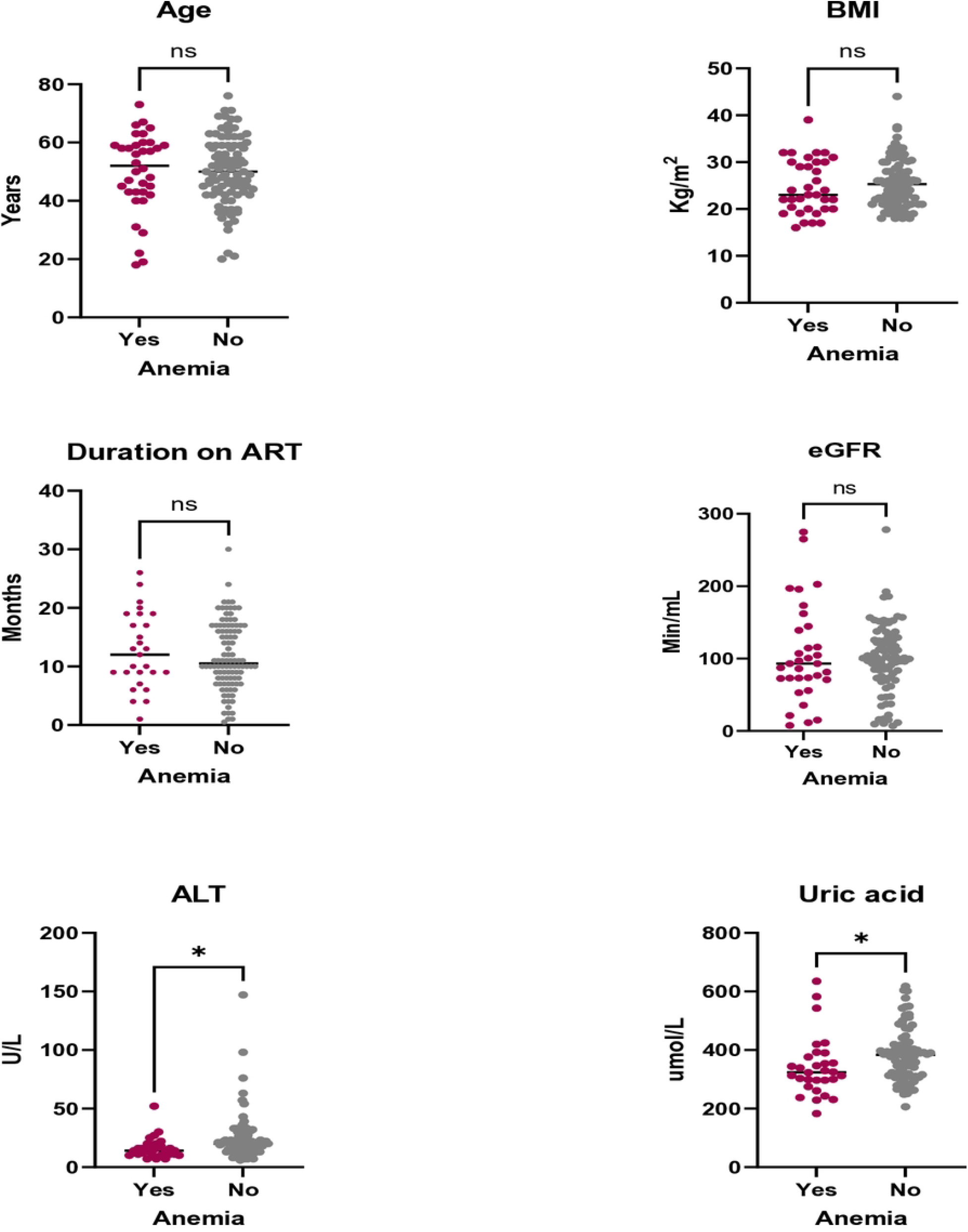
shows scatter plots comparing clinical parameters (age, BMI, duration on ART, eGFR, ALT, and uric acid levels) between HIV-positive individuals with and without anemia, with significant differences indicated for ALT and uric acid (* p < 0.05), and non-significant differences indicated for age, BMI, duration on ART, and eGFR (ns).

### Immune and Inflammatory Cytokines by Anaemic status PLWH

We examined the levels of inflammatory cytokines in virally suppressed PLWH with and without anemia (Figure 2). The findings revealed that interferon-gamma (INF-gamma) was elevated in anaemic individuals compared to their non-anaemic counterparts (P<0.001). This distinction suggests a notable variation in immune activation between the two groups, with higher INF-gamma levels observed in those experiencing anemia. In contrast, other inflammatory cytokines including TNF (tumor necrosis factor), IL-6 (interleukin-6), IL-1, IL-5, and IL-17 showed no significant differences between the anaemic and non-anaemic groups.

**Figure 2.**
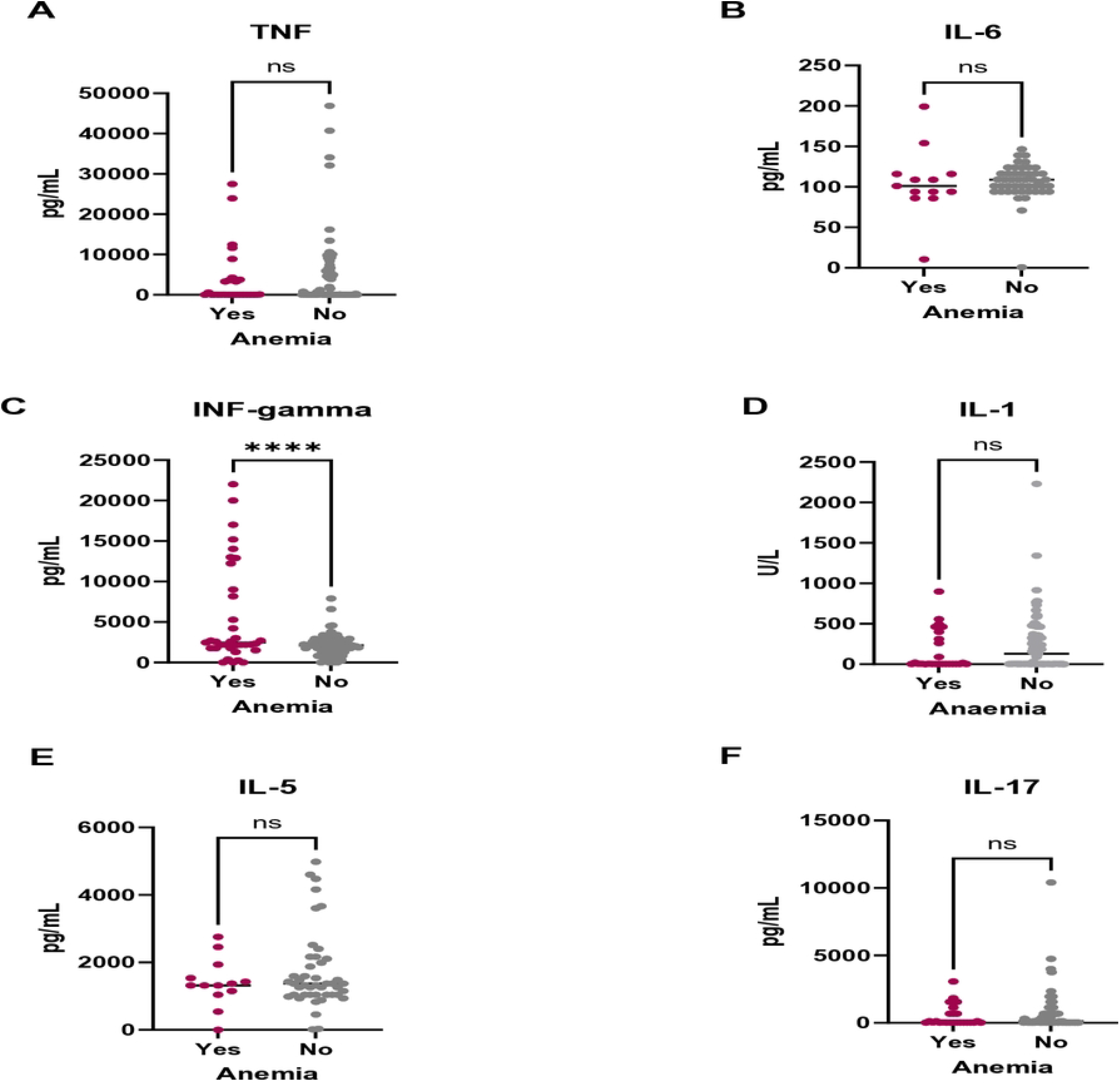
Comparison of inflammatory cytokine levels ((A)TNF, (B) IL-6, (C) INF-gamma, (D) IL-1, (E) IL-5, and (F) IL-17) between anaemic and non-anaemic virally suppressed PLWH.

### Hematologic Profiles in Anaemic and Non-Anaemic PLWH

The Figure 3 presents hematologic and iron metabolism profiles among virally suppressed PLWH, comparing individuals with and without anemia. Statistically significant differences were observed for ferritin and erythropoietin levels. Participants with anemia had significantly lower ferritin levels (p < 0.001), indicating reduced iron stores, while erythropoietin levels were significantly higher (p < 0.05), suggesting a compensatory response to anemia. No significant differences were found in soluble transferrin receptor 1 (sTFR1), hepcidin, or D-dimer levels, as denoted by ‘ns’ (not significant).

**Figure 3.**
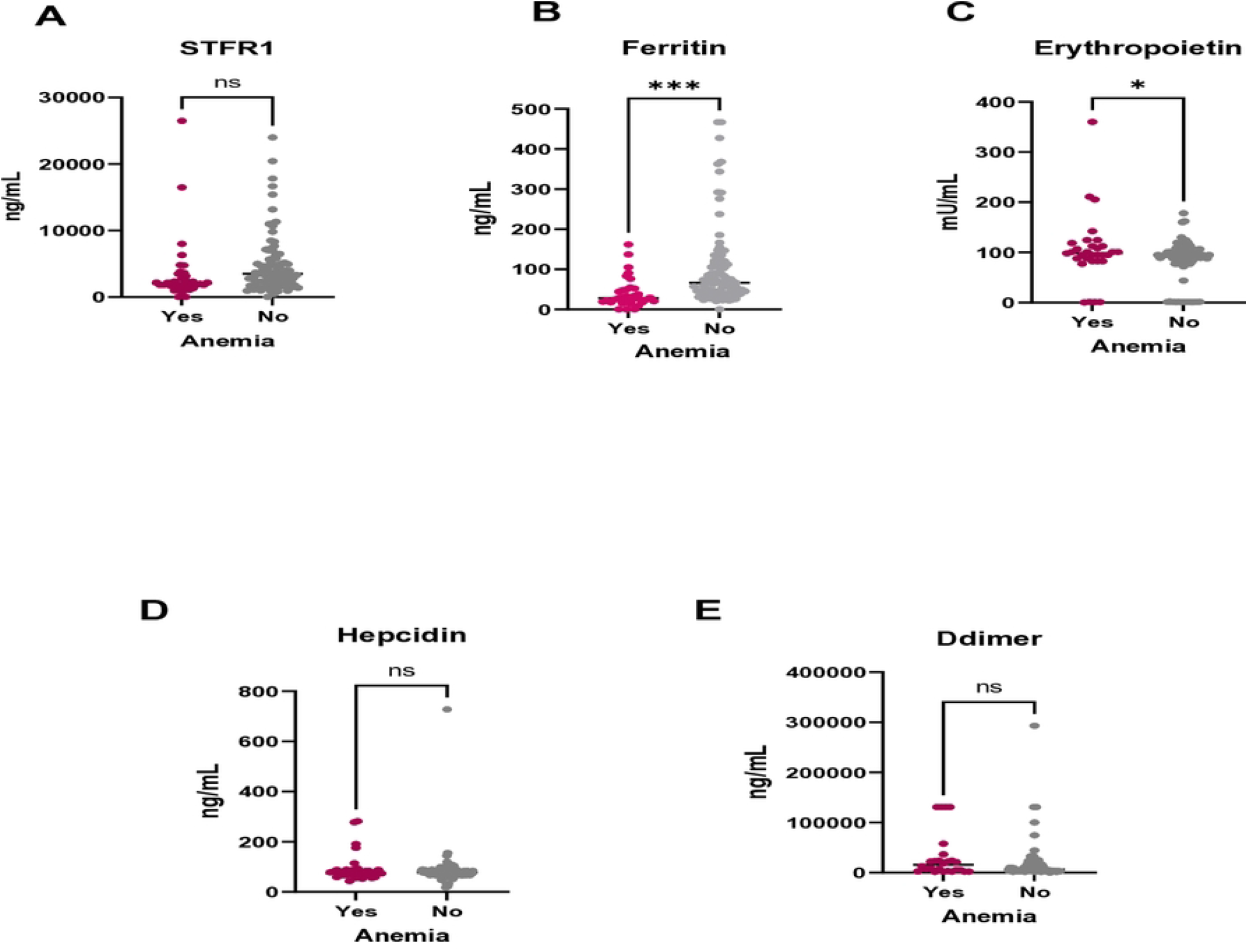
Scatter plots comparing hematologic profiles ((A) sTfR1, (B) Ferritin, (C) Erythropoietin, (D) Hepcidin, and (E) Ddimer) between individuals with and without anemia. Data points represent individual measurements, with median values indicated by horizontal lines. Statistical significance was denoted by asterisks (*** for p < 0.001) and “ns” for not significant.

### Logistic Regression Analysis of Immune Profiles in Anemia Among Virally Suppressed PLWH

In the **unadjusted analysis (Table 2)**, female sex was significantly associated with higher odds of anemia (OR = 3.09, 95% CI: 1.09-8.77, p = 0.033). Among the inflammatory markers, high-sensitivity C-reactive protein (hs-CRP) shows no association with anemia (OR = 1.00, p = 0.039), and IL-17 displays a borderline inverse association (OR = 0.99, p = 0.004). Interferon-gamma, IL-6, TNF-α, IL-1, IL-5, neutrophil count, lymphocyte count, monocyte count, SII, and NLR did not show significant associations with anemia in this unadjusted model.

**Table 2.** Immune profiles associated with anemia PLWH.

| Variables | Unadjusted analysis |  | P | Adjusted analysis |  | P |
| --- | --- | --- | --- | --- | --- | --- |
|  | OR | 95% CI |  | AOR | 95% CI |  |
| Age | 1.03 | 0.99-1.06 | 0.157 | 1.13 | 1.021-1.252 | **0.018** |
| Sex |  |  |  |  |  |  |
| Male |  |  |  |  |  |  |
| Female | 3.09 | 1.09- 8.77 | **0.033** | 3.95 | 0.23-68.76 | 0.347 |
| D-dimer *pg/mL* | 1.00 | 1.00-1.00 | 0.568 | 1.00 | 1.00-1.00 | 0.467 |
| Hs CRP *pg/mL* | 1.00 | 1.00-1.00 | **0.039** | 1.00 | 1.00-1.00 | 0.272 |
| Interferon_gamma *pg/mL* | 1.00 | 0.99-1.00 | 0.176 | 1.003 | 1.001-1.005 | **0.012** |
| Interleukin-17a *pg/mL* | 0.99 | 0.99-1.00 | **0.004** | 0.99 | 0.99-1.00 | 0.051 |
| Interleukin-6 *pg/mL* | 1.00 | 0.98-1.02 | 0.959 | 1.02 | 0.95-1.08 | 0.634 |
| Tumor Necrosis fator-α *pg/mL* | 1.00 | 1.00-1.00 | 0.113 | 1.00 | 1.00-1.00 | 0.212 |
| Interleukin-1 *U/L* | 1.00 | 1.00-1.00 | 0.718 | 1.004 | 0.99-1.01 | 0.174 |
| Interleukin-5 *pg/mL* | 1.00 | 0.99-1.00 | 0.295 | 0.99 | 0.99-1.00 | 0.13 |
| Neutrophil count *cells/L* | 1.03 | 0.66-1.62 | 0.886 | 1.65 | 0.26-10.51 | 0.595 |
| Lymphocyte count *cells* | 0.99 | 0.99-1.01 | 0.603 | 0.99 | 0.98-1.02 | 0.812 |
| Monocyte count *cells/L* | 2.15 | 0.38-12.04 | 0.385 | 3.74 | 0.11-126.22 | 0.463 |
| Systemic Immune Inflammation | 0.99 | 0.99-1.01 | 0.724 | 1.02 | 0.99-1.05 | 0.066 |
| Neutrophil Lymphocyte Ratio | 0.74 | 0.19-2.94 | 0.667 | 0.01 | 0.01-2.63 | 0.104 |
| Abbreviations: IL- Interleukins, CRP C -reactive protein, TNF Tumor necrosis factor, SII Systemic Immune-Inflammation Index, NLR Neutrophil-to-Lymphocyte Ratio, OR odds ratio, AOR Adjusted odds ratio, CI Confidence interval |  |  |  |  |  |  |

In the **adjusted analysis (Table 2)**, age was significantly associated with anemia, with each additional year associated with a 13% increase in odds (AOR = 1.13, 95% CI: 1.021-1.252, p=0.018). Female sex, however, loses significance in the adjusted model (AOR = 3.95, 95% CI: 0.23-68.76, p = 0.347). Interferon-gamma emerges as a significant factor, with slightly elevated levels linked to higher odds of anemia (AOR = 1.003, 95% CI: 1.001-1.005, p = 0.012). IL-17a maintains a borderline association with anemia (AOR = 0.99, p = 0.051), while hs-CRP and other markers, including IL-6, TNF-α, and blood cell counts, remain non-significant. SII approaches significance (AOR = 1.02, p = 0.066),

### Logistic regression analysis of hematologic profiles in virally suppressed PLHIV

In the unadjusted analysis, female participants had significantly higher odds of anemia compared to males, with an odds ratio (OR) of 3.10 (95% CI: 1.09–8.77, P=0.033). Other markers, including age, the soluble transferrin receptor-to-ferritin (STFr-Ferritin ratio), hepcidin, erythropoietin, D-dimer, MCV, and platelet count, did not demonstrate significant relationships with anemia in this model.

In the **adjusted analysis (Table 3)**, sex shows a significant association with anemia, with females experiencing a substantially higher odds (AOR = 10.01, 95% CI: 1.08-92.64, p = 0.042). An elevated sTfR1-Ferritin ratio gained significance in the adjusted model, with an AOR of 6.54 (95% CI: 1.001-42.76, p = 0.049), indicating that this biomarker may be an important factor associated with anemia. However, other variables, including age, hepcidin, erythropoietin, D-dimer, MCV, and platelet count, remain non-significant in the adjusted analysis. Overall, female sex and sTfR1-Ferritin index stand out as significant factors associated with anemia, while most other hematologic and inflammatory profiles do not show notable associations in this adjusted model.

**Table 3.** Haematologic profiles associated with anemia PLWH.

| Variables | Unadjusted analysis |  | *P* | Adjusted Analysis |  | *P* |
| --- | --- | --- | --- | --- | --- | --- |
|  | OR | 95% CI |  | AOR | 95% CI |  |
| Age | 1.03 | 0.99-1.06 | 0.157 | 1.03 | 0.97-1.09 | 0.34 |
| Sex |  |  |  |  |  |  |
| *Male* |  | 1 |  |  | 1 |  |
| *Female* | 3.10 | 1.09-8.77 | **0.033** | 10.01 | 1.08-92.64 | **0.042** |
| Hepcidin *ng/mL* | 0.99 | 0.97-1.00 | 0.530 | 0.99 | 0.99-1.01 | 0.330 |
| Erythropoietin *mU/mL* | 0.99 | 0.98-1.00 | 0.055 | 0.99 | 0.98-1.01 | 0.520 |
| STFr-Ferritin Index |  |  |  |  |  |  |
| <1 |  | 1 |  |  | 1 |  |
| 1-2 | 0.79 | 0.28-2.30 | 0.678 | 1.73 | 0.42-7.19 | 0.450 |
| >2 | 2.68 | 0.78-9.16 | 0.117 | 6.54 | 1.001-42.76 | **0.048** |
| D-dimer *pg/mL* | 1.00 | 1.00-1.00 | 0.568 | 0.99 | 0.99-1.00 | 0.940 |
| MCV *fL* | 1.21 | 0.56-2.65 | 0.627 | 2.25 | 0.96-5.27 | 0.063 |
| Platelet count | 1.002 | 0.99-1.01 | 0.523 | 1.01 | 0.99-1.013 | 0.210 |
| Abbreviations: OR odds ratio, AOR adjusted odds ratio, STFR Soluble transferrin receptor, RBC red blood cells |  |  |  |  |  |  |

## DISCUSSION

The findings in this study highlight the relationship between immune activation, iron metabolism, and anemia in virally suppressed PLWH. Advancing age was a significant factor, which may be due to the natural decline in hematopoietic function and an increased burden of comorbidities (19). In older individuals, there could often be an age-related increase in inflammation and reduced erythropoiesis, both of which can contribute to the development of anemia (20). This was consistent with existing literature, where aging has been associated with a higher prevalence of anemia in PLWH, possibly due to diminished bone marrow response and the effects of long-term HIV infection and ART on hematologic health (21).

Immune activation, particularly through interferon-gamma (IFN-γ), was associated with anemia among virally suppressed PLWH. Immune activation in virally suppressed PLWH can persist due to several factors including residual viral antigens or immune responses to low levels of viral replication in sanctuary sites such as the gut or lymph nodes (22–24). Although ART effectively reduces viral replication, it may not completely eliminate latent reservoirs, leading to low-level viremia and continuous immune stimulation. This persistent immune activation can contribute to systemic inflammation, impaired erythropoiesis, and the development or persistence of anemia in PLWH (25). These results align with prior studies, which demonstrate that IFN-γ suppresses bone marrow function, and skews HSC differentiation toward myeloid lineages, ultimately leading to HSC exhaustion (26–28). In addition the systemic immune-inflammation index (SII), which approached significance in the analysis, further emphasizes the role of systemic inflammation in anemia despite viral suppression. ART significantly reduces systemic inflammation and immune activation, but it does not bring these levels down to those observed in HIV-uninfected populations (29) (30).

The sTfR1-ferritin ratio, a marker of functional iron deficiency, was another critical component in understanding anemia in PLWH. The elevated ratio found in the study reflects a situation where iron stores are depleted in the body (15). Studies suggest that the sTfR1-ferritin ratio is a more accurate indicator of pure iron deficiency, highlighting its utility in diagnosing iron-related anemia in PLWH (8,15,31).

Other studies have reported that females generally have lower iron stores than men, making them more susceptible to iron deficiency anemia emphasizing the importance of routine screening for iron deficiency in PLWH (32). Iron deficiency in PLWH can result from multiple mechanisms but in resource-limited settings with high burden of malnutrition may be a significant contributor.

However, this study had some limitations. Potential confounders such as dietary intake, micronutrient levels (e.g., folate, vitamin B12), and undiagnosed comorbid conditions were not assessed, which could have influenced biomarker levels and anemia outcomes.

The clinical implications of these findings are significant for the management of anemia in virally suppressed PLWH. The association between immune activation, particularly elevated interferon-gamma levels, and anemia suggests that routine viral suppression alone may not be sufficient to resolve anemia in this population. Furthermore, these findings emphasize the need for closer monitoring of anemia, with the sTFR1-ferritin index recommended for improved diagnosis. This study could guide more targeted interventions, including anti-inflammatory strategies, iron supplementation, or nutritional support, depending on the underlying cause.

## Conclusion

Low grade immune activation persisted in virally suppressed individuals, as evidenced by elevated levels of interferon-gamma and an increased soluble transferrin receptor-ferritin index reflecting iron deficiency anemia due to depleted iron stores. These findings emphasize the need for enhanced anemia management strategies that account for both demographic and treatment-related factors. Monitoring immune-hematologic profiles was essential for guiding targeted interventions. Integrating these insights into clinical practice could improve early detection, management, and overall patient outcomes in PLWH, particularly those at higher risk for anemia.

## Data Availability

The data underlying the results presented in the study are available along with this submission

## Author Contributions

K.K., S.K.M conceptualized the study and K.K., wrote the draft manuscript. B.M.H., and SM edited different sections of the manuscript. K.K, SKM, B.M.H, S.M analysed the data and created all the tables and figures, S.M. and S.K.M. finalized the manuscript. All authors have read and agreed to the published version of the manuscript.

## Data availability

The datasets used during the current study is available from the corresponding author on reasonable request.

## References

1. Chirimuta LAM, Shamu T, Chimbetete C, Part C. Incidence and risk factors of anemia among people on antiretroviral therapy in Harare. South Afr J HIV Med. 2024 Aug 30;25(1):7.

2. Cao G, Wang Y, Wu Y, Jing W, Liu J, Liu M. Prevalence of anemia among people living with HIV: A systematic review and meta-analysis. EClinicalMedicine. 2022 Jan 26;44:101283.

3. Lang R, John Gill M, Coburn SB, Grossman J, Gebo KA, Horberg MA, et al. The changing prevalence of anemia and risk factors in people with HIV in North America who have initiated ART, 2007–2017. AIDS Lond Engl. 2023 Feb 1;37(2):287–98.

4. Morceau F, Dicato M, Diederich M. Pro-Inflammatory Cytokine-Mediated Anemia: Regarding Molecular Mechanisms of Erythropoiesis. Mediators Inflamm. 2010 Mar 1;2009:e405016.

5. Nemeth E, Ganz T. Hepcidin and Iron in Health and Disease. Annu Rev Med. 2023 Jan 27;74:261–77.

6. Ghaffari S. Oxidative Stress in the Regulation of Normal and Neoplastic Hematopoiesis. Antioxid Redox Signal. 2008 Nov;10(11):1923–40.

7. Obeagu EI, Obeagu GU, Ukibe NR, Oyebadejo SA. Anemia, iron, and HIV: decoding the interconnected pathways: A review. Medicine (Baltimore). 2024 Jan 12;103(2):e36937.

8. Infusino I, Braga F, Dolci A, Panteghini M. Soluble Transferrin Receptor (sTfR) and sTfR/log Ferritin Index for the Diagnosis of Iron-Deficiency Anemia A Meta-Analysis. Am J Clin Pathol. 2012 Nov 1;138(5):642–9.

9. Klisic A, Kavaric N, Kotur J, Ninic A. Serum soluble transferrin receptor levels are independently associated with homeostasis model assessment of insulin resistance in adolescent girls. Arch Med Sci AMS. 2021 May 10;19(4):987–94.

10. El-Gendy FM, El-Hawy MA, Rizk MS, El-Hefnawy SM, Mahmoud MZ. Value of Soluble Transferrin Receptors and sTfR/log Ferritin in the Diagnosis of Iron Deficiency Accompanied by Acute Infection. Indian J Hematol Blood Transfus. 2018 Jan;34(1):104–9.

11. Kamvuma K, Masenga S, Hamooya B, Chanda W, Munsaka S. Prevalence and factors associated with moderate-to-severe anemia among virally suppressed people with HIV at a tertiary hospital in Zambia. PLOS ONE. 2024 Aug 26;19(8):e0303734.

12. August-2022-Zambia-Consolidated-Guidelines.pdf [Internet]. [cited 2025 Feb 13]. Available from: https://www.differentiatedservicedelivery.org/wp-content/uploads/August-2022-Zambia-Consolidated-Guidelines.pdf?utm_source=chatgpt.com

13. WHO W. Haemoglobin concentrations for the diagnosis of anemia and assessment of severity [Internet]. 2024 [cited 2024 Apr 11]. Available from: https://www.who.int/publications-detail-redirect/WHO-NMH-NHD-MNM-11.1

14. Margetic S, Topic E, Ruzic DF, Kvaternik M. Soluble transferrin receptor and transferrin receptor-ferritin index in iron deficiency anemia and anemia in rheumatoid arthritis. Clin Chem Lab Med. 2005;43(3):326–31.

15. Oustamanolakis P, Koutroubakis IE. Soluble transferrin receptor-ferritin index is the most efficient marker for the diagnosis of iron deficiency anemia in patients with IBD. Inflamm Bowel Dis. 2011 Dec 1;17(12):E158–9.

16. Obirikorang C, Issahaku RG, Osakunor DNM, Osei-Yeboah J. Anemia and Iron Homeostasis in a Cohort of HIV-Infected Patients: A Cross-Sectional Study in Ghana. AIDS Res Treat. 2016 Mar 22;2016:e1623094.

17. Sullivan KM, Dean A, Soe MM. OpenEpi: a web-based epidemiologic and statistical calculator for public health. Public Health Rep Wash DC 1974. 2009;124(3):471–4.

18. Thottuvelil SR, Chacko M, Warrier AR, Nair MP, Rajappan AK. Comparison of neutrophil-to-lymphocyte ratio (NLR), platelet-to-lymphocyte ratio (PLR), and systemic immune-inflammation index (SII) as marker of adverse prognosis in patients with infective endocarditis. Indian Heart J. 2023 Nov 1;75(6):465–8.

19. Artz AS. Biologic vs physiologic age in the transplant candidate. Hematology. 2016 Dec 2;2016(1):99–105.

20. Wacka E, Nicikowski J, Jarmuzek P, Zembron-Lacny A. Anemia and Its Connections to Inflammation in Older Adults: A Review. J Clin Med. 2024 Jan;13(7):2049.

21. Herd CL, Mellet J, Mashingaidze T, Durandt C, Pepper MS. Consequences of HIV infection in the bone marrow niche. Front Immunol. 2023 Jul 11;14:1163012.

22. de Jong PR, González-Navajas JM, Jansen NJG. The digestive tract as the origin of systemic inflammation. Crit Care. 2016 Oct 18;20(1):279.

23. Martinez-Picado J, Deeks SG. Persistent HIV-1 replication during antiretroviral therapy. Curr Opin HIV AIDS. 2016 Jul;11(4):417–23.

24. Massanella M, Fromentin R, Chomont N. Residual inflammation and viral reservoirs: Alliance against an HIV cure. Curr Opin HIV AIDS. 2016 Mar;11(2):234–41.

25. Murray AJ, Kwon KJ, Farber DL, Siliciano RF. The Latent Reservoir for HIV-1: How Immunologic Memory and Clonal Expansion Contribute to HIV-1 Persistence. J Immunol Baltim Md 1950. 2016 Jul 15;197(2):407–17.

26. de Bruin AM, Voermans C, Nolte MA. Impact of interferon-γ on hematopoiesis. Blood. 2014 Oct 16;124(16):2479–86.

27. Morales-Mantilla DE, King KY. The Role of Interferon-Gamma in Hematopoietic Stem Cell Development, Homeostasis, and Disease. Curr Stem Cell Rep. 2018 Sep 1;4(3):264–71.

28. Pagani A, Nai A, Silvestri L, Camaschella C. Hepcidin and Anemia: A Tight Relationship. Front Physiol [Internet]. 2019 Oct 9 [cited 2024 Nov 11];10. Available from: https://www.frontiersin.org/journals/physiology/articles/10.3389/fphys.2019.01294/full

29. Hileman CO, Funderburg NT. Inflammation, Immune Activation, and Antiretroviral Therapy in HIV. Curr HIV/AIDS Rep. 2017 Jun 1;14(3):93–100.

30. Quiros-Roldan E, Castelli F, Lanza P, Pezzoli C, Vezzoli M, Biasiotto G, et al. The impact of antiretroviral therapy on iron homeostasis and inflammation profilesin HIV-infected patients with mild anemia. J Transl Med. 2017 Dec 19;15:256.

31. Dignass A, Farrag K, Stein J. Limitations of Serum Ferritin in Diagnosing Iron Deficiency in Inflammatory Conditions. Int J Chronic Dis. 2018 Mar 18;2018:9394060.

32. Abioye AI, Sudfeld CR, Hughes MD, Aboud S, Muhihi A, Ulenga N, et al. Iron status among HIV-infected adults during the first year of antiretroviral therapy in Tanzania. HIV Med. 2023;24(4):398–410.

